# Predicting the Impact of the COVID-19 Pandemic for the Low- and Middle-Income Countries

**DOI:** 10.1101/2020.08.13.20167064

**Authors:** Zhiwei Ding, Feng Sha, Jing Kong, Bingyu Li, Yi Zhang, Paul Yip, Zhouwang Yang

## Abstract

This study predicts the maximum hospital demand and number of infections for the LMICs in the first wave of COVID-19 pandemic. The epidemic is estimated to impose health care burden excessively exceeding the current capacity of hospitals in many LMICs, especially in Honduras, Central African Republic and Colombia.

---

Although most of the high-income countries have reached the peak of the first wave of COVID-19 epidemic in April and May, 2020^1^, the epidemic has just started in most of the low- and middle-income countries (LMICs). Unlike the high-income countries, the LMICs are more likely to encounter serious shortage of health care resources. Researchers have found association between mortality and health care resources, emphasizing the importance of effective disease control in resource-limited regions^2^. This study predicts the maximum hospital demand and number of infections for the LMICs in the first wave of COVID-19 pandemic, with the aim of providing specific disease control guidance for the LMICs.

The study used data on confirmed COVID-19 patients, recovered patients and deaths before July 31, 2020 from Johns Hopkins University^3^. Based on the time-varying SIR dynamic model^4^ and constrained weighted least squares method, a novel differential equation model was derived to predict the accumulated confirmed patients at the end of first wave COVID-19 pandemic and maximum hospital beds demand for each LMIC (Appendix). COVID-19 confirmed rate (per 100,000 population) was also calculated by dividing accumulated confirmed patients by total population for each LMIC. Regression model was used to predict hospital beds supply in 2020 for each LMIC (Appendix). Maximum hospital bed utilization rate was calculated by dividing maximum hospital bed demand by total hospital bed. All analysis was performed in R (version 3.6.2).

42 LMICs were included in our analysis (figure 1). At the peak of the epidemic, Honduras (499.24%, 95% CI: 375.44%~671.14%), Central African Republic (214.17%, 72.55%~761.87%) and Colombia (203.29%%, CI: 132.88%~318.09%) will have the highest hospital bed utilization rates. Of all the 42 LMICs, 40.48 % will have more than half of hospital beds occupied by COVID-19 patients. At the end of the first wave, it is estimated that South Africa (696119, 95% CI: 556958~873474), Colombia (584725, 95% CI: 428510~824511) and Indonesia (148029, 95% CI: 113416~ 194073) will have the highest accumulated confirmed patients among all the LMICs. Specifically, Armenia will finally report the highest COVID-19 confirmed rate (per 100,000 population) with 1572 (CI: 1022~2401). The confirmed rate will also be high in South Africa (1174, CI: 939~1473) and Colombia (1149, CI: 842~1620). 45.24% of the 42 LMICs are estimated to have confirmed rate of more than 100 per 100,000 population.

**Figure 1.**
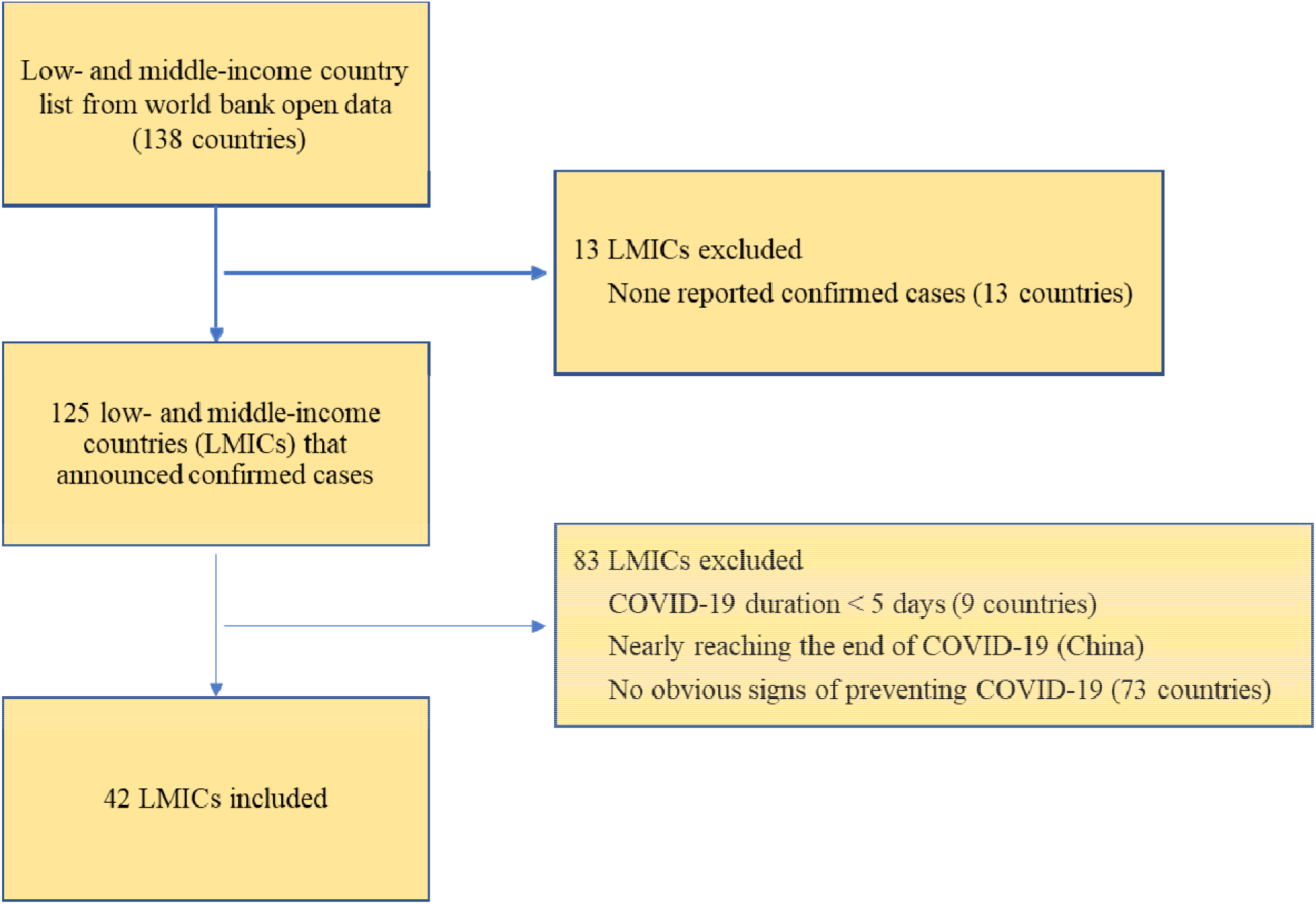
Flow chart of selecting low- and middle-income countries (LMICs) and model structure

The epidemic is estimated to impose health care burden excessively exceeding the current capacity of hospitals in many LMICs. Effective non-pharmaceutical public health interventions, including cordons sanitaire, traffic restriction, social distancing, home confinement, centralized quarantine, and universal symptom survey, should be implemented to control the spread the COVID-19^6^. In the case of uncontrollable outbreak, short-term strategies including reducing non-COVID-19 demand for health services and building mobile hospital to temporally mitigate the hospital system overload should also be recommended^5^. However, limited health care budgets in these regions may hinder the implementation of these strategies. Economic and medical assistance from more developed countries, WHO and other international NGOs will be much required for effective disease control in the LMICs.

**Table 1.** Estimated Infections, Infection Rate, Maximum Hospital Demand and Hospital Bed Utilization rate for LMICs

| Country | Number of Infections | Infection Rate<br>(per 100,000 in the country) | Maximum Hospital Demand | Maximum Hospital Bed<br>Utilization rate |
| --- | --- | --- | --- | --- |
| South Africa | 696119 (556958~873474) | 1174 (939~1473) | 216560 (155870~299353) | 161.53% (116.26%~223.29%) |
| Colombia | 584725 (428510~824511) | 1149 (842~1620) | 143221 (93618~224103) | 203.29% (132.88%~318.09%) |
| Indonesia | 148029 (113416~194073) | 54 (41~71) | 31725 (21263~46897) | 10.23% (6.86%~15.13%) |
| Honduras | 65673 (51293~85352) | 663 (518~862) | 47832 (35971~64302) | 499.24% (375.44%~671.14%) |
| Kyrgyzstan | 58213 (49715~68406) | 892 (762~1048) | 28358 (22608~35518) | 75.70% (60.35%~94.82%) |
| Uzbekistan | 50398 (32025~82968) | 151 (96~248) | 24124 (13019~45508) | 29.70% (16.03%~56.02%) |
| Armenia | 46594 (30270~71136) | 1572 (1022~2401) | 13126 (6733~24509) | 74.44% (38.18%~139.00%) |
| Azerbaijan | 41209 (36865~46049) | 406 (364~454) | 11970 (10111~14133) | 27.14% (22.92%~32.04%) |
| Bosnia and Herzegovina | 24284 (15872~38763) | 740 (484~1182) | 12539 (7127~22494) | 96.87% (55.06%~173.78%) |
| Cameroon | 23383 (8602~67118) | 88 (32~253) | 5804 (1400~24593) | 21.95% (5.29%~93.00%) |
| Cote d'Ivoire | 21131 (14954~29936) | 80 (57~113) | 9090 (5433~14845) | 121.17% (72.42%~197.88%) |
| Bulgaria | 21070 (13343~34678) | 303 (192~499) | 8444 (4450~16244) | 12.52% (6.60%~24.08%) |
| North Macedonia | 18103 (10330~32930) | 869 (496~1581) | 7882 (3504~17441) | 86.28% (38.36%~190.93%) |
| Madagascar | 14995 (9036~25760) | 54 (33~93) | 4364 (2107~9186) | 103.17% (49.81%~217.16%) |
| West Bank and Gaza | 14167 (7166~32909) | - | 8980 (3766~24733) | - |
| Central African Republic | 11032 (4183~36212) | 228 (87~750) | 9464 (3206~33667) | 214.17% (72.55%~761.87%) |
| Zambia | 9399 (2679~89633) | 51 (15~488) | 3092 (677~45261) | 11.36% (2.49%~166.33%) |
| Democratic Republic of<br>the Congo | 9337 (5719~16748) | 10 (6~19) | 4381 (2431~9438) | 9.68% (5.37%~20.85%) |
| Paraguay | 8513 (3979~19860) | 119 (56~278) | 3003 (979~9206) | 36.47% (11.89%~111.80%) |
| Haiti | 8249 (4716~31511) | 72 (41~276) | 3803 (3259~14147) | 52.24% (44.77%~194.33%) |
| Gabon | 8167 (3265~21108) | 367 (147~948) | 2250 (559~8343) | 113.98% (28.32%~422.64%) |
| Mauritania | 6282 (4647~8461) | 135 (100~182) | 2319 (1468~3597) | 186.27% (117.91%~288.92%) |
| Malawi | 6181 (2459~30297) | 32 (13~158) | 3069 (1095~20599) | 15.78% (5.63%~105.90%) |
| Zimbabwe | 5528 (1708~33146) | 37 (11~223) | 3665 (846~26678) | 16.72% (3.86%~121.71%) |
| Nicaragua | 5256 (3208~10041) | 79 (48~152) | 1351 (827~3010) | 24.85% (15.21%~55.36%) |
| Montenegro | 4479 (1695~17257) | 713 (270~2748) | 3352 (928~15545) | 142.46% (39.44%~660.65%) |
| Cabo Verde | 3157 (1777~5774) | 568 (320~1039) | 1435 (629~3224) | 138.65% (60.77%~311.50%) |
| Mali | 3111 (176~34991) | 15 (1~173) | 899 (37~21605) | 59.73% (2.46%~1435.55%) |
| Yemen | 2807 (789~10989) | 9 (3~37) | 1097 (168~6596) | 4.98% (0.76%~29.95%) |
| Namibia | 2775 (1420~7813) | 109 (56~307) | 2500 (1242~7344) | 44.49% (22.10%~130.70%) |
| Benin | 2504 (1355~6472) | 21 (11~53) | 1363 (640~4566) | 29.63% (13.91%~99.26%) |
| Guinea-Bissau | 2491 (1415~7534) | 127 (72~383) | 1729 (1044~5774) | 116.51% (70.35%~389.08%) |
| South Sudan | 2491 (1822~4270) | 22 (16~38) | 1676 (1352~2844) | - |
| Angola | 1802 (712~12832) | 5 (2~39) | 1193 (415~11294) | 7.67% (2.67%~72.64%) |
| Liberia | 1720 (730~4363) | 34 (14~86) | 841 (241~2803) | 27.02% (7.74%~90.04%) |
| Lesotho | 1021 (671~1702) | 48 (31~79) | 610 (372~1095) | 23.58% (14.38%~42.33%) |
| Burundi | 592 (177~9404) | 5 (1~79) | 105 (39~3037) | 1.33% (0.50%~38.57%) |
| Eritrea | 480 (239~3860) | 14 (7~109) | 283 (159~3324) | 12.58% (7.07%~147.73%) |
| Sao Tome and Principe | 351 (231~7849) | 160 (105~3581) | 199 (199~1837) | 37.20% (37.20%~343.36%) |
| Cambodia | 327 (171~87905) | 2 (1~526) | 132 (38~87451) | 1.06% (0.31%~704.28%) |
| Belize | 60 (44~128) | 15 (11~32) | 26 (16~85) | 6.97% (4.29%~22.79%) |
| Grenada | 26 (22~42) | 23 (20~37) | 8 (8~13) | 1.99% (1.99%~3.23%) |

*Note:*
*a. Ultimate Confirmed cases (per 100,000 in the country) = 100,000 × (Ultimate Confirmed cases / total population of the country).*
*b. Peak Bed Utilization rate = Beds Used at Peak / National Hospital Bed capacity*
*c. The list of low- and middle-income countries was obtained from World Bank Open Data. The hospital bed density data by country came from World Health Organization. The Annual Total Population across countries came from United Nations.*

## Data Availability

All the data are public available.

https://coronavirus.jhu.edu/map.html

## Acknowledgments

**Feng Sha:** Conceptualization, Validation, Resources, Writing - Original Draft. **Zhiwei Ding:** Methodology, Software, Formal analysis, Writing - Original Draft, Visualization. **Jing Kong:** Methodology, Writing - Review & Editing. **Bingyu Li:** Writing - Original Draft, Writing - Review & Editing. **Yi Zhang:** Methodology, Project administration. **Paul Yip:** Writing - Review & Editing. **Zhouwang Yang:** Conceptualization, Methodology, Resources, Writing - Review & Editing, Supervision, Funding acquisition.

This study is funded by Strategic Priority CAS Project (grant XDB38040200), National Natural Science Foundation of China (grant 71950011, 11871447, 71991464/71991460) and National Key Research and Development Program of China (grant 2018AAA0101001).

